# One Health genomics of *Acinetobacter baumannii* reveals sector-specific lineages and permeable ecological barriers

**DOI:** 10.64898/2026.04.09.26350516

**Authors:** Julie Plantade, Corentin Escobar, Anne-Sophie Godeux, Ludovic Poiré, Amédée André, Valentin Deromelaere, Pierre Cassier, Jean-Philippe Rasigade, Sylvie Nazaret, Charles Coluzzi, Samuel Venner, Maria-Halima Laaberki, Xavier Charpentier

## Abstract

*Acinetobacter baumannii* is a major cause of severe hospital-acquired infections, with a steadily increasing global prevalence driven by a few clinically adapted lineages. Animals and natural environments also harbor *A. baumannii* populations, but assessing their connections to clinical lineages is limited by sparse genomic data and a lack of integrated sampling. We conducted a local One Health genomic epidemiology study, sampling, isolating, sequencing, and characterizing several hundred *A. baumannii* isolates from clinical, animal, and environmental contexts. Within a geographically restricted area, we recovered several globally distributed clinical lineages (international clones, ICs), as well as livestock- and environment-associated lineages shared across Europe, highlighting widespread dissemination beyond clinical settings. Isolates closely related to the emerging clinical lineage IC11 were found in livestock, but no other clinically associated lineages were detected outside clinical contexts. Among these, the epidemic superlineage IC2 was identified in both human and veterinary clinical settings, indicating that similar practices in human and animal medicine select for closely related opportunistic pathogens. We found that hospitals host distinct, antibiotic-sensitive endemic populations capable of causing infection. These populations belong to a diversifying clade spanning clinical and environmental contexts and carry a high load of insertion sequences. Strong plasmid conservation further suggests frequent horizontal gene transfer across ecological compartments. Overall, *A. baumannii* comprises diverse, context-adapted lineages with a high potential for global spread. Although intercontext transmission appears limited, plasmids may overcome these ecological barriers. Our findings underscore the need for integrated One Health surveillance to better understand transmission pathways and limit the emergence of clinically adapted strains.

## Introduction

*Acinetobacter baumannii* is an opportunistic pathogen primarily associated with hospital-acquired infections. Historically, its distribution has been largely confined to the human clinical sector, where it has established a series of globally disseminated high-risk lineages known as International Clones (IC1–IC11) (1). Among these, genomic analyses have consistently revealed the predominance of IC2, and its main representative sequence type 2 (ST2), which has become the most successful and widespread lineage worldwide (1). A key feature of *A. baumannii* is its remarkable ability to acquire resistance to multiple classes of antibiotics, including carbapenems, which are often considered drugs of last resort (2). Carbapenem resistance is primarily mediated by the acquisition of carbapenem-hydrolyzing class D β-lactamases such as OXA-23 (encoded by *bla*_OXA-23_), often embedded in a variety of transposons that facilitate horizontal gene transfer (3). Less commonly, resistance arises from metallo-β-lactamases such as New Delhi metallo-β-lactamase (NDM), which confers the ability to hydrolyze almost all β-lactams, including carbapenems (4). Another mechanism involves OXA-58 (*bla*_OXA-58_ gene), which typically exhibits low-level carbapenem hydrolysis but can confer higher resistance when overexpressed through insertion sequence (IS)-mediated upregulation (4). These resistance determinants can be chromosomally encoded or plasmid-borne, promoting their persistence and dissemination across bacterial populations. Recently, attention has turned to the emergence of the IC11 lineage (ST164), first described in 2014 (5, 6) and increasingly reported across different geographic regions. Although its prevalence remains low, IC11 is of particular concern as early isolates carried the *bla*_OXA-23_ gene, while more recent representatives have instead acquired *bla*_NDM-1_ (7).

The prevalence of international clones in human medicine has overshadowed the presence of *A. baumannii* in other sectors. In companion animals, several studies have reported infections and nosocomial outbreaks involving lineages, such as ST25 and ST2, usually associated with human infections (8–11). These findings are not unexpected given the close physical contact and healthcare similarities between humans and pets. *A. baumannii* has also been detected among livestock. The seasonal occurrence of *A. baumannii* in cattle raises the question about whether these detections represent transient contamination or carriage (12). Fatal cases in farm animals caused by carbapenem-resistant ST2 strains have also been reported (11). Expanding beyond animal hosts, studies have identified *A. baumannii* strains in environmental sources such as soil and plants, often associated with decomposing organic matter (13, 14). These findings collectively suggest that *A. baumannii* may have a broader ecological niche than previously appreciated. Despite these emerging data, isolates from non-human sources remain underrepresented in genomic databases. This sampling bias limits our ability to reconstruct the ecological diversity and evolutionary trajectories that underpin *A. baumannii*’s adaptation to the clinical environment. Addressing this gap is critical to understand how environmental and animal reservoirs may contribute to the emergence and dissemination of clinically relevant lineages. Adopting this integrated approach will be key to anticipating future evolutionary shifts and mitigating the spread of multidrug-resistant lineages across sectors.

In the present study, we investigated the genetic diversity of *A. baumannii* isolates collected within a geographically restricted area, encompassing human, domestic animal, livestock and environmental compartments. By combining high-resolution genomic analyses with comparative datasets from broader international collections, we aimed to identify potential transmission dynamics across distinct ecological and geographical contexts. This integrative approach provides new insights into how local populations of *A. baumannii* connect to the global genetic landscape of this pathogen.

## Material and methods

### Sampling design and isolation methods

The four hospital sites of the Hospices Civils de Lyon (HCL) were sampled from January to December 2022. About 1,200 samples were obtained from the neonatal & pediatric care wards, adult intensive care units (burn unit, surgical care unit, neurological unit), specialized medical units (hepato-gastroenterology and transplant, internal medicine, geriatric medicine, long-term care) and infectious and tropical diseases units. In total, 24 distinct surface types were randomly sampled (nurse cart, bedside rail, over-bed table, patient chair, bedside monitor, television remote control, workbench, computer, door handle, sink/faucet, telephone, laboratory coat,…). Dry surfaces were sampled using sterile non-woven wipes moistened with buffered peptone water containing 10 % neutralizer (Dominique Dutscher, Bernolsheim, France), in accordance with International Standard ISO 18593 for sampling techniques. Samples were collected before daily room cleaning (i.e. ∼24 h after the last previous daily cleaning). The wipes were incubated for 48 h at 36 °C in tryptone soy broth, which was placed into a stomacher (for 30 s, speed 2). Samples were then plated on MacConkey agar plates (Thermo Fisher Scientific). Species identification was performed with standard microbial methods (MALDI-TOF Vitek MS BioMérieux). Putative *A. baumannii* colonies were purified and stored at −80°C until further species identification (see *A. baumannii* identification section).

Veterinary clinic was sampled between April 2021 and April 2022 in the same intensive care unit (ICU) of the hospital at the veterinary medicine school of Lyon in France (SIAMU, VetAgro Sup, Marcy l’étoile). *A. baumannii* isolates retrieval was previously reported (15). In the present study, the 14 isolates were subjected to new sequencing along with isolates of other sectors (see dedicated section).

For cattle sampling, study design was approved by VetAgro Sup ethic committee approval no. 2078. Samples were taken by nasal swabbing at the feed fence during morning or evening milking (Amies Agar M40 Transystem) between May 21 and September 15, 2021. The four farms with more than one positive animal were sampled regularly between May and September 2022 (four sampling sessions of 15 cattle per farm) and then a single sampling session of 15 cattles in each farm between May 31 and June 6, 2023. Swabs were kept at room temperature in the transport medium 6 to 24 hours before plating on selective medium (CHROMagar Acinetobacter (CHROMagar) prepared according to the manufacturer’s instructions without addition of the CHROMagar MDR supplement CR102.

Soil samples were collected along a transect of a stagnant branch of the Rhône river upstream of the city of Lyon. Soil samples were placed into sterile, zip-sealed plastic bags and stored at –20°C until processing. For direct isolation, soil samples were homogenized within their collection bags prior to inoculation. A sterile cotton swab was inserted into the bag and gently rotated for several seconds to ensure even contact with the soil matrix. The swab was then streaked directly onto CHROMAgar Acinetobacter plates (CHROMAgar, France). Plates were incubated at 37°C for 24–48 h. For enrichment-based isolation, approximately 1 mg of homogenized soil was suspended in 3 mL of minimal salt medium supplemented with 0.2% acetate (KH_2_PO_4_, 73.5 mM; Na_2_HPO_4_, 35.2 mM; (NH_4_)_2_SO_4_, 15.1 mM; MgSO_4_·7H_2_O, 0.81 mM; FeSO_4_·7H_2_O, 0.004 mM; CaCl_2_·2H_2_O, 0.007 mM) in a sterile 12 mL culture tube and vortexed thoroughly. The suspension was incubated at 37°C for 5 h with agitation. Following incubation, 200 μL of the culture was plated onto CHROMAgar and incubated at 37°C for 24–48 h prior to colony selection.

### A. baumannii identification

Colonies morphologically similar to *A. baumannii*, including those previously identified by MALDI-TOF, were subjected to PCR identification using primers annealing on the *bla*_OXA-51_ gene (forward primer, OXA51WG-F, 5’-ATGAACATTAAAGCACTCTTAC; reverse primer OXA51WG-R, 5’-CTATAAAATACCTAATTGTTCT) according to previous work (16).

### Genome sequencing and assembly

Genomic DNA from bacterial samples was extracted from pellets obtained from 1 mL of exponential-phase culture using the DNeasy Blood & Tissue Kit (Qiagen, Hilden, Germany). All extracted DNA were then purified using SPRI beads (NucleoMag, Macherey-Nagel, Düren, Germany). Sequencing libraries were prepared and sequenced using the Oxford Nanopore MinION platform with R10.4 (for native barcoding libraries) or R10.4.1 (for rapid barcoding librairies) flow cells. Native barcoding libraries were obtained using the Native Barcoding Expansion Kit (SQK-NBD112.24), and rapid barcoding libraries were obtained with the Rapid Barcoding Kit (SQK-RBK004.24), following the manufacturer’s protocols. The target sequencing depth was a minimum of 100 Mb per genome. If initial coverage was insufficient, additional sequencing runs were conducted, and data were merged prior to assembly. Reads were base-called using Dorado with super accurate model (sup). Genomes were assembled with Hybracter (17) and Autocycler (18). Assemblies were inspected for complete chromosome and for the presence of plasmids that some assembler could miss. When assemblies were identical in terms of contigs, the Autocycler assembly was kept by default. Otherwise, the assembly with a complete chromosome and the most plasmids (inspected for the presence of replication genes) was selected. Checkm (1.2.3) (19) showed that completeness was above 97% and contamination was below 2% for all assemblies. Due to their lower accuracy, assemblies with coverage lower than 30x were included in the phylogeny and gene presence/absence analyses but omitted from epidemiology analyses. Genome assemblies are available under bioproject PRJNA1301220. Assembly accession numbers and metrics are listed in **Table S1**.

### Sequence typing and AMR genes identification

Sequence type Pasteur scheme (referred to ST) (20) and Oxford scheme (STOxford) (21) alleles were obtained with mlst_check (2.1.1706216, option -s ‘Acinetobacter baumannii’) and compared with PubMLST sequence type database (retrieved on 2025-06-10). Assignment for new ST of the Pasteur Scheme were submitted to PubMLST for genomes with coverage ≥50X. AMR genes identification was conducted with ResFinder (4.6.0) (22).

### Core genes maximum likelihood phylogenies

We first constructed a core genes phylogeny using our 364 isolates and *A. nosocomialis* M2 strain as an outgroup (GCF_005281455.1). Assemblies were annotated with Bakta (23). Then, pangenome at 80% of identity was obtained using PanACoTA (1.4.0) (24). Persistent genes at 0.99 prevalence were identified with PanACoTA corepers, option -t 0.99, and aligned (n genes = 2008) with PanACoTA align. The alignment was used to build a maximum likelihood phylogeny using IQTree with model GTR+F+I+G4 (identified by ModelFinder), 1000 ultrafast bootstraps (--ufboot 1000) and UFBoot tree optimization (--bnni). We also built a phylogeny with this study’s isolates, 471 isolates from the NCBI database and 5 *A. nosocomialis* strains as outgroup (M2, NIPH286: GCF_000368605.1, NIPH2119: GCF_000368085.1, TG19596: GCF_000301695.1 and TG21145: GCF_000301775.1). Public isolates were selected according to the following criteria: number of contigs < 350 and contig N50 > 45kb. All assemblies underwent syntactical annotation with prodigal implemented in PanACoTA (PanACoTA annotate with option –prodigal). Then, 80% of identity pangenome was constructed, followed by persistent genes identification (0.99 prevalence) and their alignment. Phylogeny was built with IQTree using GTR+F+I+G4 (identifies by ModelFinder), 1000 ultrafast bootstraps (--ufboot 1000) and UFBoot strees optimization (--bnni). Phylogeny visual representations were build using iTOL (7.4.2).

### Gene content correlation matrix and mash distances

Matrices of gene presence were constructed based on PanACoTA pangenomes (80% identity). Pearson’s correlation coefficients were obtained with function cor() of package stats in R. Mash distances were obtained using Mash (2.3) ‘dist’ command with default parameters (25).

### Plasmids, phages and IS detection

Plasmids and phages identification was conducted using geNomad (1.11.0) ‘end-to-end’ command with parameters –lenient-taxonomy and –full-ictv-lineage to allow for classification retrieval (26). IS were identified using ISEscan (1.7.3) with default parameters (27). Plasmid replicase families (Rep_1, Rep_3, PriCT_1) were identified in each plasmid by predicting ORFs with Prodigal (meta mode) (28) followed by HMM searches using HMMER (29) against Pfam profiles (PF01446, PF01051, PF08708) (30). Plasmid sequences were compared using all-vs-all nucleotide searches with BLAST+, and results were integrated with plasmid metadata (context) and host genomic distances estimated by Mash. Only reciprocal plasmid pairs meeting ≥95% identity and ≥95% coverage in both directions and carried by genetically distant strains (Mash distance >0.01) were retained (25).

## Results and discussion

### An intersectorial, time- and geographically-restricted sampling recovers a large and diverse *A. baumannii* population

We sought to capture the diversity of *A. baumannii* within a geographically-restricted area across ecological contexts of varying levels of anthropogenic activities. Looking for a natural context, a stagnant branch of the Rhône river upstream of city of Lyon was found positive for *A. baumannii*. At this location, soil samples were collected near nettles root or *Impatiens glandulifera* roots during the warm season (July to November) (14). Six sampling campaigns over a 3-years window (2020, 2021, 2022) yielded 102 natural context isolates (**Fig. S1**). A higher level of anthropogenic activity is represented by the livestock context. Cattles from 5 out of 8 investigated dairy farms of a rural area on the western side of Lyon were found positive for carriage of *A. baumannii*. The four farms with the highest isolation rates were sampled on a yearly basis (2021 to 2023) (**Fig. S1**), resulting in 156 livestock isolates. *A. baumannii* were also isolated from the clinical context, both in human and animal sectors. A previously reported sampling campaign conducted in 2021-2022 on cats and dogs in a veterinary intensive care unit yielded 13 hospitalized pets isolates and one veterinary clinical environment isolate (15). From January to December of 2022, all four hospital sites of the city of Lyon (north, south, east and central) were subjected to random sampling of the ward environment (furniture, coat, trolley, mobile units, waiting rooms, bed rails, sink, bench), recovering 64 human clinical environment isolates. During that same time period, 28 *A. baumannii* isolated from patients (infection and screening) were retrieved from the centralized microbiology service.

In total, within a 30 km-radius geographical area and over a 3 years period, we collected 364 isolates forming the Ab-One collection. Ab-One consists of nearly equal numbers of isolates from a natural context (n=102), livestock context (n=156) and veterinary and human clinical contexts (n=106) (**Table S1**). Complete genomes obtained by long-read sequencing revealed a core genome of 2,532 genes and a pangenome of 27,080 genes (90% identity, 95% prevalence). Comparatively, the most recent pangenome analysis using 5,824 representative genomes for *A. baumannii* identified 36,992 pan genes and 2,266 softcore genes (1). Thus, with only 364 isolates recovered from within a small geographical area (<30 km of radius), the Ab-One collection shows a pangenome size 70% of that of the species worldwide. The phylogenetic tree of the Ab-One isolates, obtained from the alignment of persistent core genes (80% identity, 99% prevalence, n genes=2008), showed an overall unbalanced topology with some closely branching lineages along with many other long branches (**Fig. 1**). Thus, our intersectorial sampling campaign recovered a large and diverse meta-population of *A. baumannii*, composed of isolates from recent clonal lineages (possibly outbreak) and several highly divergent lineages.

**Fig. 1.**
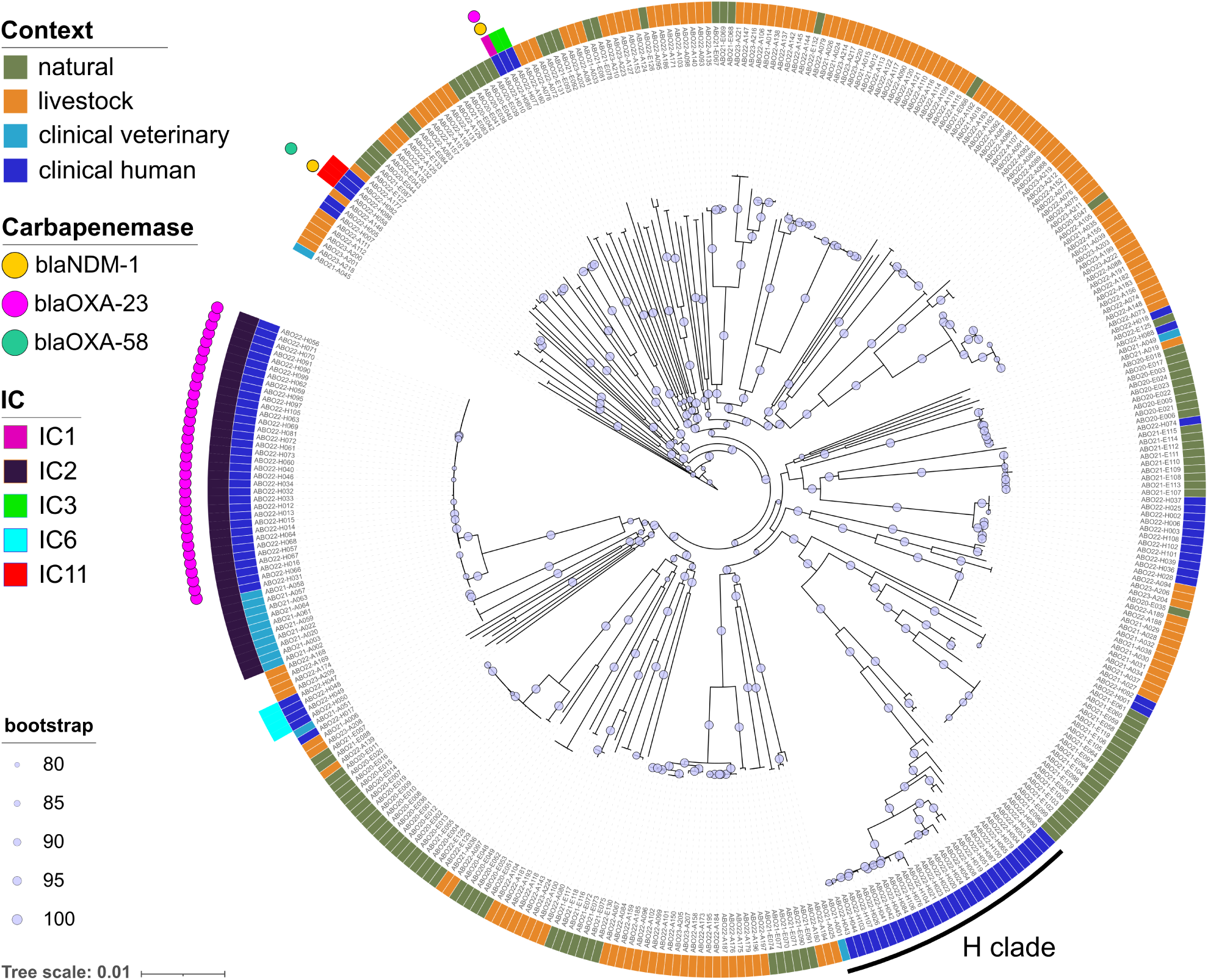
Core genome phylogeny of the 364 isolates of the Ab-One collection. Maximum likelyhood phylogeny tree from the alignment of persistent genes (99% prevalence, 80% identity). The tree is rooted using *A. nosocomialis* M2 strain as an outgroup. Inner ring: isolates are colored by the context of their isolation (clinical human, dark blue; clinical veterinary, light blue; livestock, orange; natural, green). Middle ring: international clone (IC). The outer ring correspond to carbapenemases identified in the genomes of isolates. Bootstrap values ranging from 80 to 100 are shown. Tree scale is in substitution per site.

### Hospital sites harbor specific endemic and epidemic A. baumannii populations

Isolates of the clinical context fall in at least 9 distinct lineages scattered throughout the Ab-One phylogenetic tree, representing 16 known ST and 6 new ones. However, most isolates are grouped into two distinct clades, a diversifying clade encompassing 9 STs (thereafter referred to as the H clade), and a clonal clade of isolates of ST2, the representative ST of the IC2. These ST2 isolates formed two clonal subclades. One exclusively composed of veterinary ICU epidemic isolates lacking carbapenemase (15). The other subclade includes another carbapenemase-lacking veterinary isolate which branches out of a highly clonal group of 34 CRAb isolates possessing the OXA-23 carbapenemase. All 34 isolates encode it as part of Tn*2008* (31) within a 76 kb conjugative plasmid also encoding resistance to aminoglycosides, sulfonamides and tetracyclines. Nearly identical variants of this plasmid (1–3 SNPs), consistent with horizontal spread by conjugation, have been detected in clinical ST2 strains across distinct ST^Oxford^ types : first in Pakistan in 2012 during a sheep pneumonia outbreak (ST^Oxford^ 452, CP024578.1) (11), then in the USA in 2021 (ST^Oxford^ 1144, CP069841.1), and finally in a human infection in Pakistan in 2022 (STOxford 1289, CP147864.1). All Ab-One ST2 CRAb isolates were collected in the burn ICU of the central hospital site over an 8-month time frame (January to August), from infections (n=4 isolates), patient carriage (n=6) or hospital environment (n=24), indicative of an epidemic that remained contained. We used the Mash distance of their whole genome to further estimate their relationship. Two main groups show a Mash distance of 0, one with 12 isolates sampled between January and April 2022 and another with 16 isolates sampled between April and August 2022. This suggests that at least two epidemic clones might have persisted in burn ICU between January and August 2022.

The same central hospital site hosted isolates of ST belonging to four other recognized ICs: IC1, IC3 (ST3), IC6 (ST78) and IC11 (ST164). Moreover, one CRAb isolate, ABO22-H010, is a single locus variant (*fusA*) of ST1, the representative of IC1, and was assigned the new ST3319. Sampled on furniture from the burn ICU, its genome carries resistance determinants to aminoglycosides, trimethoprim, sulfonamides, tetracyclines and carbapenems (*bla*_NDM-1_, *bla*_OXA-23_). Out of three ST164 isolates (IC11) (5), isolate ABO22-H058 is also a CRAb (*bla*_OXA-58_ and *bla*_NDM-1_), with resistance determinants to aminoglycosides, macrolides, sulfonamides and tetracyclines. All of these genes were located on a 110 kb plasmid (pABO22-H058.1). Only few descriptions of plasmids co-harboring *bla*_NDM-1_ and *bla*_OXA-58_ were made in *Acinetobacter towneri* or non-IC11 *A. baumannii* (32–34). However, pABO22-H058.1 shares no significant identities with these sequences except for their Tn*125*-like harboring *bla*_NDM-1_. Although IC11 isolates wre previously reported to carry *bla*_OXA-23_ or chromosomal *bla*_NDM-1_ on a Tn*6924*-like element, this is the first description of a plasmid carrying *bla*_OXA-58_ and *bla*_NDM-1_ genes in this emerging clone (5).

While the ST2 clade is only populated by isolates of the central site, a diversifying clade (hereafter called H clade) encompass isolates from the central, east and north sites (**Fig. 1** and **Fig. S2**). Within this H clade, three ST (ST203, ST1639 and ST2532) are found in at least two sites. Mash distance between ST203 strains of different sites vary from 0.0019 to 0.0037, arguing against clonality and recent intersite transfer. Similar results were obtained between isolates of three different sites within ST1639 (**Fig. S2**), with the exception of ABO22-H045 (east) and ABO22-H084 (north) showing a mash distance of 4.8×10^-4^. Gene content analysis (r=0.97) indicated difference in their accessory genomes (**Fig. S2**). Indeed, ABO22-H084 harbors five IS insertions (three IS3, one IS630, one IS5), a 17 kb IS5-based transposon, a prophage region and a 6-kb plasmid that were absent in H045. Conversely, three IS insertions (IS3, IS5, IS6), two small indels, two prophages and two plasmids (approximately 3 kb and 6 kb) were specific to H045. These many genomic divergence are not consistent with a recent clonal spread between hospital sites. Similar mash distance was observed between ST2532 isolates of the north site and east sites, and, outside of the H clade, between two ST3 isolates from the east and central sites. Outside of the H clade, only another ST (ST32) is found across three sites (east, north and south), with again no evidence of clonal spread. Hence, there is limited evidence of intersite transfer, consistent with east and north site showing diverse isolates of non overlapping ST.

The H clade also contains a veterinary isolate, ABO21-A001, sampled in an animal ICU in May 2021 (15). This isolate clusters with human isolates of ST2532 (**Fig. S2**), however Mash distances (0.00053 to 0.0015) and gene content did not suggest clonality (**Fig. S2**). These observations suggest that ST2532 isolates might be adapted to both human and veterinary clinic, potentially sharing genetic factors other than antibiotic resistance (resistance to desiccation, etc.).

Given the high prevalence of ST2, we assessed its association with infection. We analyzed the dependence between STs and the source of samples. Four isolates sampled in infected patients were ST2 and 7 were non-ST2. Thirty isolates sampled during routine screening or in hospital environment were ST2 and 51 were non-ST2. Exact Fisher test on the contingency table showed that ST2 was not more frequently associated with an infection (p-value = 1, odds ratio = 0.972, 95%IC[0.192, 4.206]), despite including data from an ST2 local epidemic.

Overall, sampling four hospital sites in the course of 2022, highlighted the presence of international clones 1, 2, 3, 6 and 11. ST2 CRAb and other non-ST2 CRAb were restricted to a single hospital site hosting the burn patients ICU. Beside the high prevalence of ST2 in the hospital site hosting the burn patient ICU, each hospital site showed an endemic, diverse and antibiotic-susceptible *A. baumannii* population. The recurrence of some STs suggest their adaptation to the human and veterinary clinical context.

### Persistent populations of *A. baumannii* in farm animals

In the livestock context, *A. baumannii* isolates (n = 156) exhibited greater genetic diversity than those from the clinical setting, with 34 distinct ST identified, including 19 newly described lineages. This high diversity among cattle isolates is consistent with findings reported in Lebanon, France, and Germany (12, 35, 36). No resistance genes were detected in any Ab-One farm isolates following screening against comprehensive resistance gene databases. This genomic profile strongly suggests that these isolates remain susceptible to antibiotics such as ampicillin combined with β-lactamase inhibitors or to cephalosporins. This presumed high susceptibility contrasts with the phenotypic resistance previously reported in cattle isolates in Germany, where all isolates (n = 126) were resistant to ampicillin–clavulanic acid, cephalexin, and ceftiofur (12). Regional differences in antibiotic usage practices or farming systems may contribute to this discrepancy, particularly given the high usage of third-generation cephalosporins such as ceftiofur in dairy cattle reported in the investigated German region.

Phylogenetic analysis revealed that farm-associated strains did not form a single monophyletic group but were instead distributed across multiple clades (**Fig. 1**). To assess whether specific lineages were recurrently associated with cattle farms, we examined the temporal and spatial distribution of ST (**Fig. 2A**). Among the most prevalent lineages, four ST (935, 1027, 1013, and 927) were repeatedly detected within the same farms over the two-year sampling period and were also shared between two or three different farms.

**Fig. 2.**
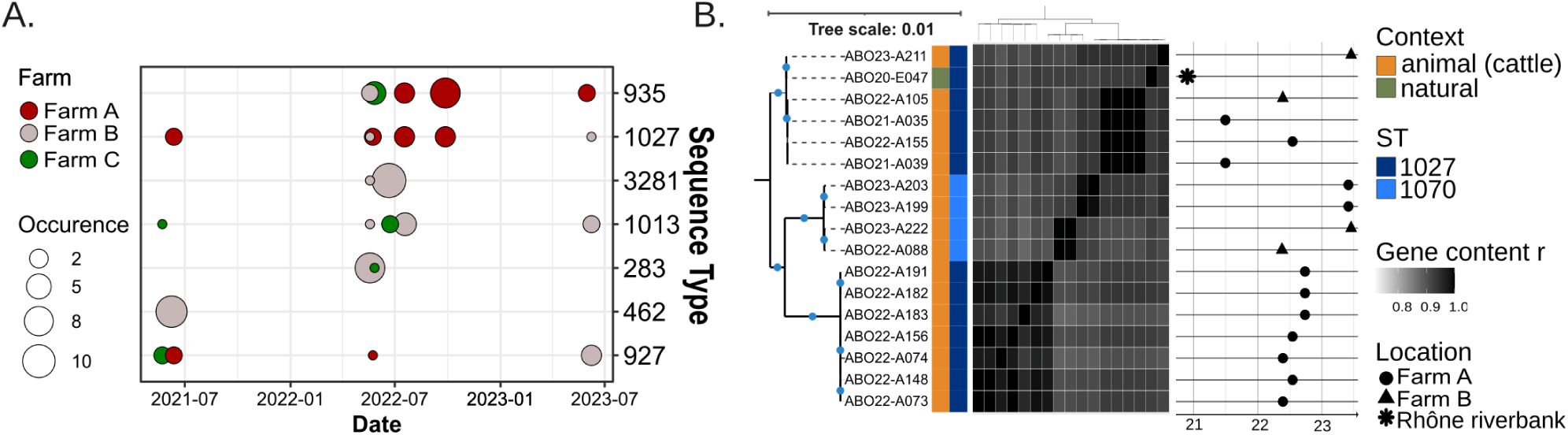
*A. baumannii* lineages associated with cattle. Panel (A) shows the distribution over time and across farms of the seven most abundant *A. baumannii* ST lineages detected in the livestock context (ordered top to bottom). Each bubble represents a detection event, with bubble size proportional to the number of isolates. Colors indicate the farm of origin. Panel (B), core-genome phylogeny and gene content similarity of cattle farm isolates belonging to the ST1027–1070 lineage. The phylogenetic tree (left; scale = 0.01 substitutions/site) shows clusters supported by bootstrap values ≥ 80 (blue dots). Adjacent color bars indicate sampling context and ST type. The central heatmap displays pairwise gene-content similarity (r). To the right, the timeline indicates the sampling date of each isolate, with symbols denoting sampling location.

Closer examination of the phylogenetic relationships among isolates belonging to these shared ST confirmed the clustering of isolates originating from the same farm and sampling date (**Fig. 2B** and **Fig. S4**). This pattern strongly supports the hypothesis of contamination from a common source within individual farms. However, a notable exception was observed within the ST1027–1070 clade (**Fig. 2B**), where isolates from different farms (ABO21-A035 from farm A and ABO22-A105 from farm B) clustered closely together and show a Mash distance of 0. These findings suggest potential cross-farm transmission over a distance of approximately 20 km (**Fig. S1**), possibly involving clones adapted to persist in these environments.

### High diversity of *A. baumannii* populations in the natural environment

Altogether, the isolates from the natural context fall into 31 ST, 8 of which are newly identified. In agreement with their low anthropized context, the isolates have genomes that do not encode detectable AMR genes. One exception being isolates of the ST354, which carry the tet(39) gene on a 9.8 kb plasmid and encoding a tetracycline efflux transporter. Isolates of the natural context are scattered throughout the tree and tend to be on long branches. Yet, a clade encompasses 5 ST (106, 145, 239, 203 and 354) and appears to share a common ancestor with the clinical H clade (**Fig. 1**). Interestingly, although isolates of some ST could be found in different samples, they are almost always from the same sampling date. Only ST203 an ST2816 were found in two distinct collection dates. Hence, each sampling campaign tends to reveal a previously unseen lineage. These observations were intriguing, considering that all isolates were retrieved over the course of 3 years in one restricted location. These observations suggest that local *A. baumannii* populations are highly diverse, but that the relative advantage of individual lineages may fluctuate over time according to environmental conditions, such as temperature, rainfall, or soil humidity. This could reflect a form of temporal niche partitioning in the exploitation of shared resources, potentially stabilizing their coexistence and maintaining local diversity. Alternatively, the site may experience substantial population turnover, possibly driven by episodic influxes of distinct lineages through water flow or other environmental inputs. Despite the absence of recurrence during the sampling campaign, several STs were nevertheless previously found in other soil samples outside France: ST536 on a plant root (Germany, 2017), ST1301 (Germany, 2017), ST2816 (Nigeria) and ST155 (Germany 2020). These observations suggest that some STs might be widely distributed and associated to soil in natural environment.

### Clinical, livestock and natural contexts overlap and are shared with other European countries

To assess potential overlap between *A. baumannii* populations from clinical, animal (livestock and wildlife), and natural contexts, we expanded our genome dataset with strains collected from humans (n = 111), animals (wild or domesticated, n = 270), and natural environments (soil and plant roots, n = 90) derived from two published datasets (13, 14, 37) (**Table S2**). Incorporating these 471 additional genomes, we reconstructed a core-genome phylogeny (**Fig. S3**), that revealed limited but detectable genetic connections between clinical and livestock/wild animal isolates, and more frequent overlaps between animal and environmental isolates.

Notably, isolates belonging to ST935 were recurrently detected in three cattle farms (Fig. 2A) but also, strikingly, in a human infection case from Germany (2019), and—more expectedly—in wild animals such as the white stork and shrew (**Fig. S5**; mash distance 0.0015–0.003). These findings indicate occasional overlaps between clinical and animal populations of *A. baumannii*. With regard to IC proximity between clinical and non-clinical isolates, IC11 isolates of four ST164 from Lyon were closely related to ST241—a single-locus variant of ST164—identified in two domestic (cattle, Lyon and Canada) and one wild animal (white stork, Poland) isolates (**Fig. 3A**). However, additional genomes from both clinical and non-clinical origins are needed to refine the evolutionary history of this recently emerged global clone (5, 6).

**Fig. 3.**
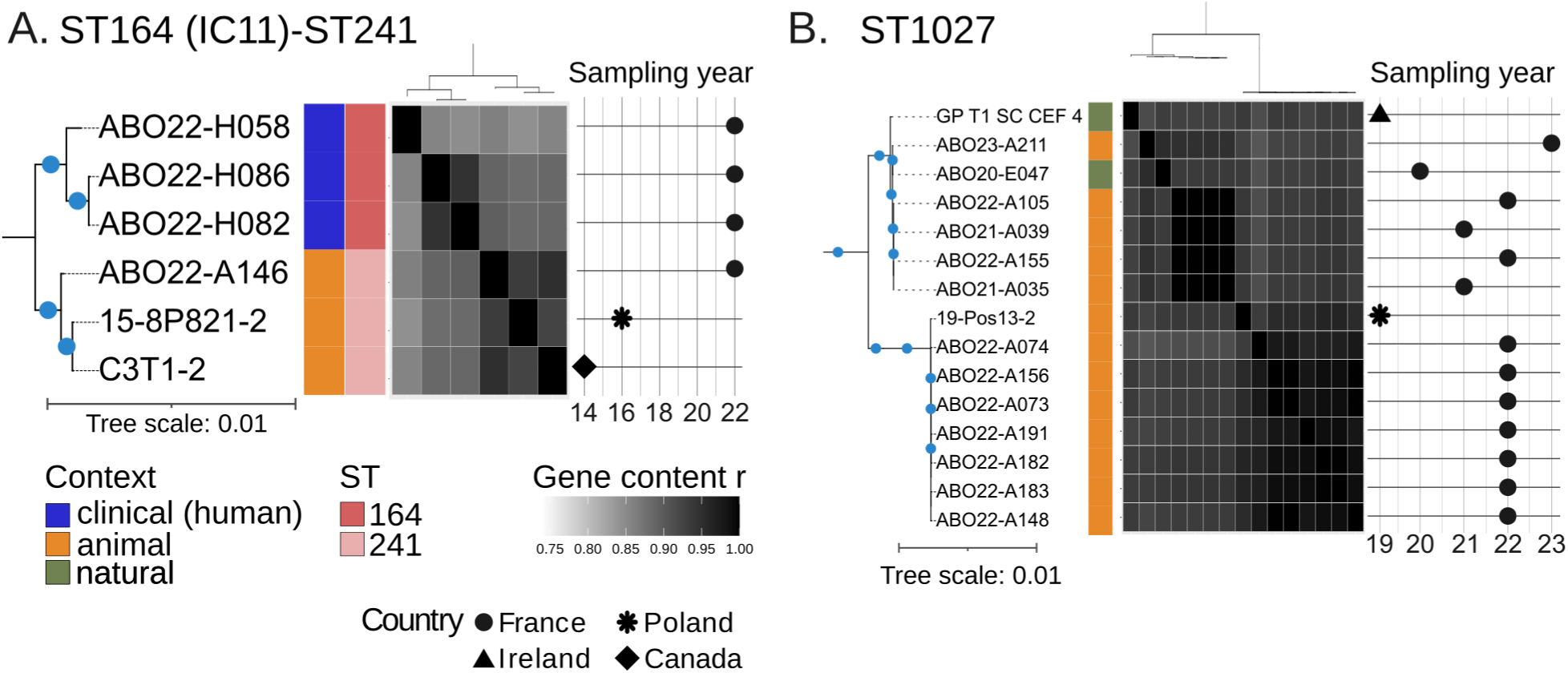
*A. baumannii* lineages shared between divers ecological and geographical contexts. Core-genome phylogenies and gene content similarity of isolates from Ab-One and additional genome sets belonging to the ST164/241 (A) and ST1027 lineages (B). The phylogenetic tree (left; scale = 0.01 substitutions/site) shows clusters supported by bootstrap values ≥ 80 (blue dots). Adjacent color bars indicate sampling context and ST type (from left to right resp.). The central heatmap displays pairwise gene-content similarity (r). To the right, the timeline indicates the sampling year of each isolate, with symbols denoting country of sampling.

Phylogenetic analysis revealed more frequent and closer relationships among isolates from animals (livestock and wildlife) and environmental sources across Europe. Persistent clones found across up to three farms of the Lyon’s region (ST935, 1027, 1013 and 927) were also identified in animals, mainly white storks, in other European countries (Poland and Germany). These isolates cluster together in the extended phylogeny, and display high gene content similarity (**Fig. 3B**. and **Fig. S5**). Moreover, Ab-One farm isolates from ST1027 are closely related to isolates from soil or grass from the same geographic region (ABO21-E047) but also from Ireland (13). Similarly, Ab-One farm isolates of the ST1013 lineages share high gene content similarity with telluric isolates from Germany and Poland (**Fig. S5**). These findings suggest continuous contamination of cattle from a common environmental reservoir (e.g., grass or soil). However, we cannot rule out the possibility that grass- or soil-adapted clones have established a transient colonization cycle in cattle, potentially facilitated by airborne transmission within the barn environment. Yet, this latter hypothesis is not consistent with the seasonal isolation pattern in cattle (12). Similarly, isolates of the ST155 isolated punctually in one farm (C in year 2022) are closely related to three Rhône riverbank isolates. The complete genomes of Ab-One offered some clues about the source of difference in the accessory genomes of closely-related isolates from different contexts. For instance, based on core-genome alignment ST155 ABO21-E067 and ABO22-A135 (natural and livestock contexts, respectively) appeared clonal, contrasting with a Mash distance of 0.001. Comparative analysis revealed that the isolates essentially differ by their prophages and plasmid content. ABO22-A135 possesses two prophages absent in isolate E067, while isolate E067 carries three distinct prophages absent from ABO22-A135. Each isolate carries a distinct 8 kb plasmid. Thus, aside from these mobile genetic elements, the two isolates would effectively be considered clonal. Similar situations are observed with ST1027 harboring an isolate of natural context (ABO20-E047) within livestock isolates (**Fig. 3B**).

Overall, our results indicate that *A. baumannii* isolates from hospitals exhibit fewer genetic connections with those from animal and environmental sources. Indeed, few lineages were shared between clinical and animal isolates, and direct links between clinical and environmental isolates were comparatively rare. In contrast, strong and recurrent connections were observed between *A. baumannii* populations from animals – both wild and domesticated – and natural environments, suggesting the existence of shared ecological lineages. Remarkably, isolates originating from different habitats, countries, and sampling years displayed high genetic proximity, raising questions about the mechanisms driving *A. baumannii* persistence and dissemination across disparate environments. Our observations also support the role of mobile genetic elements in shaping this species ecology.

### Intersectionality of mobile genetic elements at a local scale

The complete genomes of the collection allowing for an exhaustive census, we investigated the MGEs load in the Ab-One collection. Clinical isolates stand out by carrying more MGE (prophages, plasmids and IS) than isolates from livestock or natural contexts (**Fig. 4A**, **4B** and **Fig. S6**). Clinical isolates carrying twice as many prophages (up to 12) than isolates from the natural context, while livestock isolates fall in between (**Fig. S6**). This trend is similar to analyses performed on more than 4,000 prophages from diverse STs and geographical origins revealing more prophages associated with clinical context than with natural context (38). This suggests that, counter-intuitively, *A. baumannii* faces lower pressure from viral predation in the natural environment than in the clinical setting. Alternatively, the higher number of prophages in clinical isolates might protect them against lytic phages. Interestingly, three cattle farm isolates and two riverbank isolates showed no detectable prophages.

**Fig. 4.**
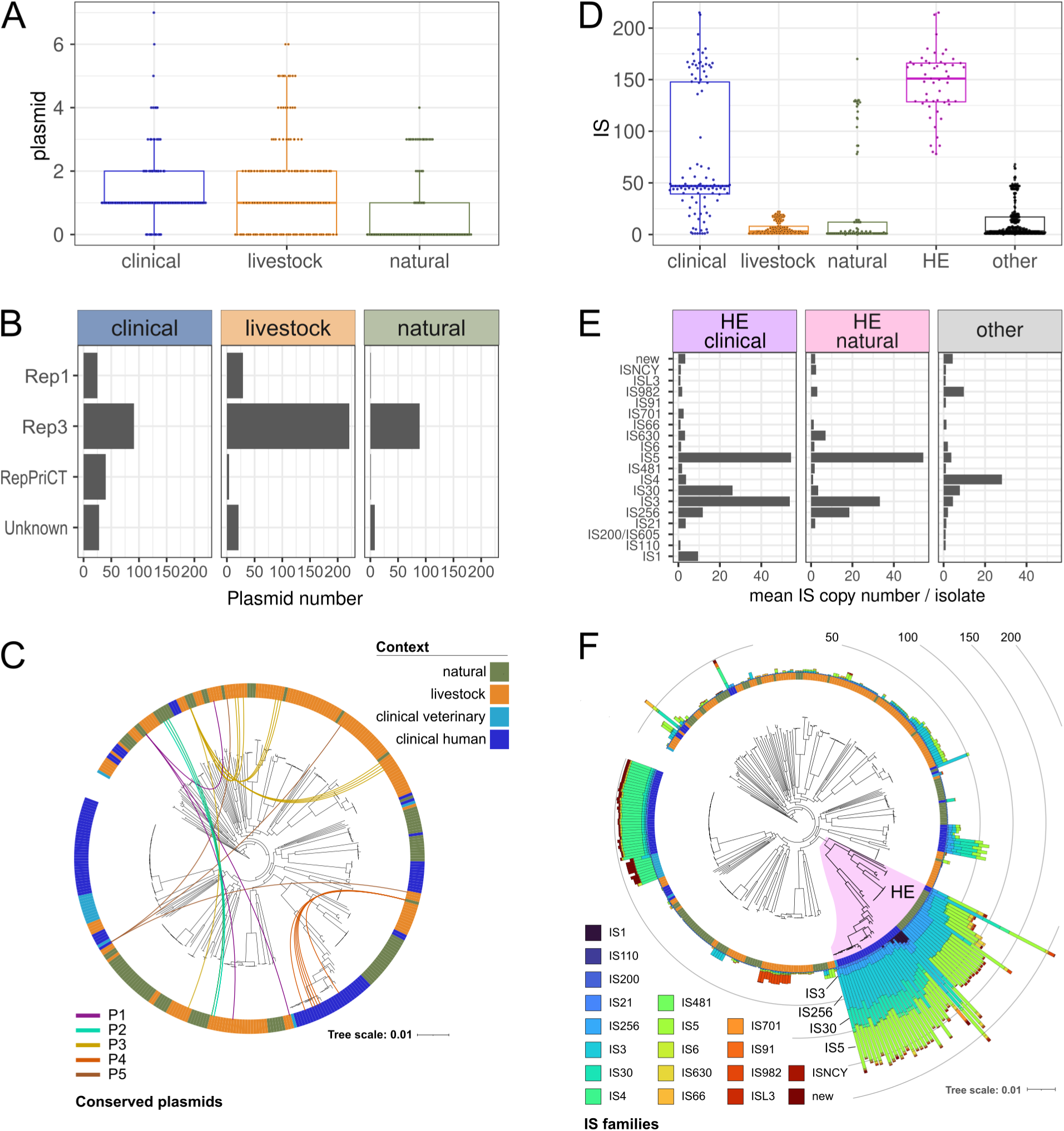
Distribution of mobile genetic elements across isolates of the Ab-One collection as a function of the ecological contexts. Panels (A) show the number of plasmids detected per genome and panel (B) the number of plasmids per context categorized based on their replicase type (Rep1, Rep3 or RepPriCT). Panel (C) represents core-genome phylogenetic tree of Fig. 1 with connecting lines indicating 5 conserved plasmids (P1 to P5 are plasmids with >95% identity and >95% coverage) between distant isolates (Mash>0.01) from distinct contexts. Panel (D) shows the number of insertion sequence (IS) detected per genome and (E) shows the mean IS copy number per isolate across major IS families in the clinical and natural isolats of the HE clade compared to the rest of the collection (other). Panel (F) represents core-genome phylogenetic tree of Fig. 1 with the number of IS per genome, colored by IS family and annotated by ecological context. The HE clade is highlighted in purple.

Plasmids are uncommon in isolates of the natural context, with only 31% percent of them carrying from 1 to 4 plasmids (**Fig. 4A**). Plasmid load is higher in livestock isolates, with more isolates carrying plasmids (70%) and in higher number, between 1 and 6. Clinical isolates carry the highest load with 90% of them carrying between 1 and 7 plasmids. This latter figure echoes previous studies, based on public genomes of mostly clinical source, that found that 80% of *A. baumannii* strains carry plasmids with the majority carrying only one plasmid (39). We further characterized the plasmids of the Ab-One collection (n= 558) by typing their replicase. The plasmid repertoire of *A. baumannii* identified three major groups based on replication initiation protein (Rep) phylogeny: RepPriCT, Rep1, and, Rep3 (39–41). RepPriCT plasmids are large conjugative plasmids that carry adaptive genes such as antibiotic resistance genes. Rep1 plasmids are small, with very few genes that were a minority and so far only found in the clinical ST1 lineage (40). Plasmids of the Rep3 replicase family, with an “iteron-type” replication system, were found the most abundant in *A. baumannii* genomes. Indeed, we found Rep3 plasmids to be predominant in our collection (n=400), however, this was followed by Rep1 (n=55) then RepPriCT (n=45), whereas 58 could not be typed. The extended diversity of the Ab-One collection reveals that Rep1 are not restricted to the ST1 clinical isolates and are abundant in livestock isolates (n=29) but also in non-IC clinical strains (n=25) (**Fig. 4C**). As previously suggested and as expected, RepPriCT large conjugative plasmids were mainly encountered in clinical context. We then looked for possible cross-context horizontal plasmid transfer by comparing plasmid identity between non-clonal strains (Mash distance >0.01). We identified five distinct plasmids sharing more than 95% identity and coverage ( **Fig. 4E**) between strains of the natural and livestock contexts (**Fig. 4E**, noted P2 to P5, Rep3 plasmids) and more surprisingly between strains of the natural and clinical contexts (P1, Rep1 plasmids). This is consistent with horizontal transfer of these plasmids occuring at a local scale. Interestingly, none of these plasmids carry conjugation genes that would allow for self-transfer.

### High IS load in an intersectorial clade hints at population bottlenecks

The analysis of the load of insertion sequence (IS) shows highly contrasted IS distributions (**Fig. 4C**). Livestock consistently showed few IS (mean per isolate = 6; median = 3), with the exception of a clade within the ST935 which experienced an expansion of IS982, with up to 16 copies. Natural isolates showed more IS (mean per isolate = 24) but the distribution is bimodal (median = 1). High IS load appeared as a defining feature of clinical isolates showing and average of 74 per isolates (median = 47), yet as for natural isolates, we noted a marked bimodal distribution. We expected clinical isolates to carry around 50 IS elements. This expectation was based on a previous analysis performed on 1035 *A. baumannii* genomes, mostly from clinical isolates belonging to IC2, which found an average of 33 copies of IS elements per genome (42). The ST2 isolates from our collection are in agreement with this study, carrying for instance over 30 copies of IS4, the IS family of IS*Aba*1 constituting the Tn*2006* transposon carrying *bla*_OXA-23_. Both in natural and clinical isolates, the bimodal distribution is due to isolates belonging the HE clade, formed by the previously reported clinical H subclade and an adjacent E subclade that mostly contains natural isolates (**Fig. 4D**). While the clinical and natural isolate seemed to be respectively distributed along these two subclades, the extended phylogeny revealed wildlife and clinical isolates intermixed with natural isolates (**Fig. S3**). With a mean number of IS per isolate of 146 (median = 151), this HE clade showed the highest number of IS (mean outside the clade = 12). Although natural isolates from the HE clade displayed a number of IS (mean = 117; median = 126) lower than in the clinical isolates of HE (mean = 163; median = 164), they showed a strikingly similar IS content with an enrichment in IS5, IS3 and IS256 (**Fig. 4E**). This is consistent with the H and E subclades as having evolved from a common ancestor readily carrying some of these IS. The clinical H subclade differs in that it further experienced expansion of IS30, and of which another subclade showed a surge of IS1 insertions. The genomes of the HE clade do not show antibiotic resistance genes, and the IS elements of this clade do not carry cargo of obvious selective advantage. Thus, why is the IS load in this HE clade well above levels reported for most bacterial genomes (43, 44)? The high load is reminiscent of a small subset of species accumulating large numbers of IS (often >100), such as *Shigella* or *Yersinia pestis* (43, 45, 46). Proliferation of IS elements in these species is thought to reflect inefficient purifying selection in populations of small effective size and experiencing frequent bottlenecks, typical of bacteria with host-restricted or intracellular lifestyle (47). This suggests that isolates of the HE clade *A. baumannii*, found in hospitals, soil or wildlife have a small effective population size and cannot control IS expansion through purifying selection. IS should also be susceptible to gene deletions, a process that may be have become less effective in the HE clade (47). Regardless of the ecological and evolutionary scenario leading to this pattern, the high transposon load may by driving the diversification of this clade.

## Conclusion

Our One-Health sampling approach recovered a highly diverse population of *A. baumannii* with novel lineages indicating that the diversity of the species has not yet been captured in full, specifically in the natural and animal compartments. The recovery of globally distributed lineages (IC1, IC2, IC3, IC6, IC11) is consistent with a high potential of this opportunistic pathogen for global dissemination and one can expect that the newly lineages identified in this study are also globally distributed. In spite of their global distribution, some lineage are associated with specific ecological contexts. In hospital settings, contamination of the environment, patient carriage and infections involving the internationally recognized clone IC2 underscores its high adaptation to the clinical environment, whether human or veterinary. However, the presence of non-IC isolates specific to the clinical context suggests that other lineages are well adapted for persistence in this environment. Although these are often not resistant to antibiotics, some isolates were recovered from infected patients suggesting a potential for causing outbreaks. Livestock and natural environments exhibit population of even greater genetic diversity, with farm isolates forming persistent, farm-specific populations and natural isolates showing minimal antibiotic resistance. Notably, overlaps with environmental populations indicate potential of cross-context transmission between grass-associated population and grazing animals at the isolate level but also through plasmid transfer. Although we found little evidence of overlap between hospital and natural population, mobile genetic elements may link these populations. Besides sharing plasmids, we uncovered a diversified clade that encompasses both clinical and natural isolates. They share an exceptionally high load of IS, suggesting a role for these mobile genetic elements in driving their diversification and ecological adaptation. Given its association to both the natural and clinical context, and the role of IS in innovation and acquisition of antibiotic resistance, the HE clade should be closely monitored for the possible emergence of clinically-adapted and antibiotic-resistant lineages.

Altogether our findings emphasize the need to better characterize the *A. baumannii* populations in human, animal, and environmental contexts and on a global scale. This is essential to elucidate the ecological pathways and transmission routes that enable *A. baumannii* to emerge from its environmental reservoirs and establish in humans populations.

## Data Availability

All data produced in the present work are contained in the manuscript

https://www.ncbi.nlm.nih.gov/bioproject/PRJNA1301220/

## Acknowledgements

We thank Benjamin Youenou and Sandra Ferraro for technical assistance during the preliminary sampling campaign of environmental isolates. We thank Diane Congiu for contributing to sampling and identification of *A. baumannii* in the hospital setting.

## References

1. Li S, Jiang G, Wang S, Wang M, Wu Y, Zhang J, Liu X, Zhong L, Zhou M, Xie S, Ren Y, He P, Lou Y, Li H, Du J, Zhou Z. 2025. Emergence and global spread of a dominant multidrug-resistant clade within Acinetobacter baumannii. Nat Commun 16:2787.

2. Müller C, Reuter S, Wille J, Xanthopoulou K, Stefanik D, Grundmann H, Higgins PG, Seifert H. 2023. A global view on carbapenem-resistant Acinetobacter baumannii. mBio 14:e02260–23.

3. Nigro SJ, Hall RM. 2016. Structure and context of Acinetobacter transposons carrying the oxa23 carbapenemase gene. J Antimicrob Chemother 71:1135–1147.

4. Rodrigues DCS, Silveira MC, Pribul BR, Karam BRS, Picão RC, Kraychete GB, Pereira FM, de Lima RM, de Souza AKG, Leão RS, Marques EA, Rocha-de-Souza CM, Carvalho-Assef APD. 2024. Genomic study of Acinetobacter baumannii strains co-harboring blaOXA-58 and blaNDM-1 reveals a large multidrug-resistant plasmid encoding these carbapenemases in Brazil. Front Microbiol 15.

5. Hansen F, Porsbo LJ, Frandsen TH, Kaygisiz ANS, Roer L, Henius AE, Holzknecht BJ, Søes L, Schønning K, Røder BL, Justesen US, Østergaard C, Dzajic E, Wang M, Ank N, Higgins PG, Hasman H, Hammerum AM. 2023. Characterisation of carbapenemase-producing Acinetobacter baumannii isolates from danish patients 2014-2021: detection of a new international clone - IC11. Int J Antimicrob Agents 62:106866.

6. 2024. Multicenter retrospective genomic characterization of carbapenemase-producing Acinetobacter baumannii isolates from Jiangxi patients 2021–2022: identification of a novel international clone, IC11. mSphere 9.

7. Zafer MM, Hussein AFA, Al-Agamy MH, Radwan HH, Hamed SM. 2021. Genomic Characterization of Extensively Drug-Resistant NDM-Producing Acinetobacter baumannii Clinical Isolates With the Emergence of Novel blaADC-257. Front Microbiol 12.

8. Naing SY, Hordijk J, Duim B, Broens EM, van der Graaf-van Bloois L, Rossen JW, Robben JH, Leendertse M, Wagenaar JA, Zomer AL. 2022. Genomic Investigation of Two Acinetobacter baumannii Outbreaks in a Veterinary Intensive Care Unit in The Netherlands. Pathogens 11:123.

9. Lupo A, Châtre P, Ponsin C, Saras E, Boulouis H-J, Keck N, Haenni M, Madec J-Y. 2017. Clonal Spread of Acinetobacter baumannii Sequence Type 25 Carrying blaOXA-23 in Companion Animals in France. Antimicrob Agents Chemother 61.

10. Bednarczuk L, Chassard A, Plantade J, Charpentier X, Laaberki M-H. 2025. Phenotypic and genetic heterogeneity of Acinetobacter baumannii in the course of an animal chronic infection. Microb Genom 11:001352.

11. Linz B, Mukhtar N, Shabbir MZ, Rivera I, Ivanov YV, Tahir Z, Yaqub T, Harvill ET. 2018. Virulent Epidemic Pneumonia in Sheep Caused by the Human Pathogen Acinetobacter baumannii. Front Microbiol 9:2616.

12. Klotz P, Higgins PG, Schaubmar AR, Failing K, Leidner U, Seifert H, Scheufen S, Semmler T, Ewers C. 2019. Seasonal Occurrence and Carbapenem Susceptibility of Bovine Acinetobacter baumannii in Germany. Front Microbiol 10.

13. Mateo-Estrada V, Tyrrell C, Evans BA, Aguilar-Vera A, Drissner D, Castillo-Ramirez S, Walsh F. 2023. Acinetobacter baumannii from grass: novel but non-resistant clones. Microbial Genomics 9.

14. Wilharm G, Skiebe E, Michalska A, Higgins PG, Weber K, Schaudinn C, Neugebauer C, Görlitz K, Meimers G, Rizova Y, Blaschke U, Heider C, Cuny C, Drewes S, Heuser E, Jeske K, Jacob J, Ulrich RG, Bocheński M, Kasprzak M, Burda E, Ciepliński M, Kaługa I, Jankowiak Ł, Aguirre JI, López-García A, Höfle U, Jagiello Z, Tobółka M, Janic B, Zieliński P, Kamiński M, Frisch J, Siekiera J, Wendel AF, Brauner P, Jäckel U, Kaatz M, Müller S, Lübke-Becker A, Wieler LH, von Wachsmann J, Thrukonda L, Helal M, Epping L, Wolf SA, Semmler T, Jerzak L. 2026. Acinetobacter baumannii’s lifestyle includes soil-dwelling colonization of decaying plant material and airborne spread. Nat Commun 17:2316.

15. André A, Plantade J, Durieux I, Durieu P, Godeux A-S, Decellieres M, Pouzot-Nevoret C, Venner S, Charpentier X, Laaberki M-H. 2024. Genomics unveils country-to-country transmission between animal hospitals of a multidrug-resistant and sequence type 2 Acinetobacter baumannii clone. Microb Genom 10:001292.

16. Pournaras S, Gogou V, Giannouli M, Dimitroulia E, Dafopoulou K, Tsakris A, Zarrilli R. 2014. Single-Locus-Sequence-Based Typing of bla OXA-51-like Genes for Rapid Assignment of Acinetobacter baumannii Clinical Isolates to International Clonal Lineages. Journal of Clinical Microbiology 52:1653–1657.

17. Bouras G, Houtak G, Wick RR, Mallawaarachchi V, Roach MJ, Papudeshi B, Judd LM, Sheppard AE, Edwards RA, Vreugde S. 2024. Hybracter: enabling scalable, automated, complete and accurate bacterial genome assemblies. Microb Genom 10:001244.

18. Wick RR, Howden BP, Stinear TP. 2025. Autocycler: long-read consensus assembly for bacterial genomes. Bioinformatics btaf474.

19. Parks DH, Imelfort M, Skennerton CT, Hugenholtz P, Tyson GW. 2015. CheckM: assessing the quality of microbial genomes recovered from isolates, single cells, and metagenomes. Genome Res 25:1043–1055.

20. Diancourt L, Passet V, Nemec A, Dijkshoorn L, Brisse S. 2010. The population structure of Acinetobacter baumannii: expanding multiresistant clones from an ancestral susceptible genetic pool. PLoS One 5:e10034.

21. Bartual SG, Seifert H, Hippler C, Luzon MAD, Wisplinghoff H, Rodríguez-Valera F. 2005. Development of a multilocus sequence typing scheme for characterization of clinical isolates of Acinetobacter baumannii. J Clin Microbiol 43:4382–4390.

22. Bortolaia V, Kaas RS, Ruppe E, Roberts MC, Schwarz S, Cattoir V, Philippon A, Allesoe RL, Rebelo AR, Florensa AF, Fagelhauer L, Chakraborty T, Neumann B, Werner G, Bender JK, Stingl K, Nguyen M, Coppens J, Xavier BB, Malhotra-Kumar S, Westh H, Pinholt M, Anjum MF, Duggett NA, Kempf I, Nykäsenoja S, Olkkola S, Wieczorek K, Amaro A, Clemente L, Mossong J, Losch S, Ragimbeau C, Lund O, Aarestrup FM. 2020. ResFinder 4.0 for predictions of phenotypes from genotypes. J Antimicrob Chemother 75:3491–3500.

23. Schwengers O, Jelonek L, Dieckmann MA, Beyvers S, Blom J, Goesmann A. 2021. Bakta: rapid and standardized annotation of bacterial genomes via alignment-free sequence identification. Microbial Genomics 7:000685.

24. Perrin A, Rocha EPC. 2021. PanACoTA: a modular tool for massive microbial comparative genomics. NAR Genom Bioinform 3:lqaa106.

25. Ondov BD, Treangen TJ, Melsted P, Mallonee AB, Bergman NH, Koren S, Phillippy AM. 2016. Mash: fast genome and metagenome distance estimation using MinHash. Genome Biology 17:132.

26. Camargo AP, Roux S, Schulz F, Babinski M, Xu Y, Hu B, Chain PSG, Nayfach S, Kyrpides NC. 2024. Identification of mobile genetic elements with geNomad. Nat Biotechnol 42:1303–1312.

27. ISEScan: automated identification of insertion sequence elements in prokaryotic genomes | Bioinformatics | Oxford Academic. https://academic.oup.com/bioinformatics/article/33/21/3340/3930124?login=false. Retrieved 17 October 2025.

28. Hyatt D, Chen G-L, LoCascio PF, Land ML, Larimer FW, Hauser LJ. 2010. Prodigal: prokaryotic gene recognition and translation initiation site identification. BMC Bioinformatics 11:119.

29. Eddy SR. 2011. Accelerated Profile HMM Searches. PLOS Computational Biology 7:e1002195.

30. Mistry J, Chuguransky S, Williams L, Qureshi M, Salazar GA, Sonnhammer ELL, Tosatto SCE, Paladin L, Raj S, Richardson LJ, Finn RD, Bateman A. 2021. Pfam: The protein families database in 2021. Nucleic Acids Res 49:D412–D419.

31. Nigro S, Hall RM. 2015. Distribution of the blaOXA-23-containing transposons Tn2006 and Tn2008 in Australian carbapenem-resistant Acinetobacter baumannii isolates. J Antimicrob Chemother 70:2409–2411.

32. Li Y, Qiu Y, Fang C, Tang M, Dai X, Zhang L. 2022. Characterisation of a novel GR31 plasmid co-harbouring blaNDM-1 and blaOXA-58 in an Acinetobacter sp. isolate. J Glob Antimicrob Resist 29:212–214.

33. Alattraqchi AG, Mohd Rani F, A. Rahman NI, Ismail S, Cleary DW, Clarke SC, Yeo CC. 2021. Complete Genome Sequencing of Acinetobacter baumannii AC1633 and Acinetobacter nosocomialis AC1530 Unveils a Large Multidrug-Resistant Plasmid Encoding the NDM-1 and OXA-58 Carbapenemases. mSphere 6:10.1128/msphere.01076-20.

34. Li Y, Qiu Y, Fang C, Dai X, Zhang L. 2022. Coexistence of blaOXA-58 and blaNDM-1 on a Novel Plasmid of GR59 from an Acinetobacter towneri Isolate. Antimicrobial Agents and Chemotherapy 66:e00206–22.

35. Pailhoriès H, Belmonte O, Kempf M, Lemarié C, Cuziat J, Quinqueneau C, Ramont C, Joly-Guillou M-L, Eveillard M. 2015. Diversity of Acinetobacter baumannii strains isolated in humans, companion animals, and the environment in Reunion Island: an exploratory study. International journal of infectious diseases: IJID: official publication of the International Society for Infectious Diseases 37:64–69.

36. Al Atrouni A, Hamze M, Rafei R, Eveillard M, Joly-Guillou M-L, Kempf M. 2016. Diversity of Acinetobacter species isolated from different environments in Lebanon: a nationwide study. Future Microbiol 11:1147–1156.

37. Mazzamurro F, Chirakadavil JB, Durieux I, Poiré L, Plantade J, Ginevra C, Jarraud S, Wilharm G, Charpentier X, P C Rocha E. 2024. Intragenomic conflicts with plasmids and chromosomal mobile genetic elements drive the evolution of natural transformation within species. PLoS Biol 22:e3002814.

38. Tenorio-Carnalla K, Aguilar-Vera A, Hernández-Alvarez AJ, López-Leal G, Mateo-Estrada V, Santamaria RI, Castillo-Ramírez S. 2024. Host population structure and species resolution reveal prophage transmission dynamics. mBio 15:e02377–24.

39. Lam MMC, Hamidian M. 2024. Examining the role of Acinetobacter baumannii plasmid types in disseminating antimicrobial resistance. npj Antimicrob Resist 2:1.

40. Lam MMC, Koong J, Holt KE, Hall RM, Hamidian M. 2022. Detection and Typing of Plasmids in Acinetobacter baumannii Using rep Genes Encoding Replication Initiation Proteins. Microbiology Spectrum 11:e02478–22.

41. Castro-Jaimes S, Guerrero G, Bello-López E, Cevallos MA. 2022. Replication initiator proteins of Acinetobacter baumannii plasmids: An update note. Plasmid 119–120:102616.

42. Adams MD, Bishop B, Wright MS. 2016. Quantitative assessment of insertion sequence impact on bacterial genome architecture. Microb Genom 2:e000062.

43. Wagner A, Lewis C, Bichsel M. 2007. A survey of bacterial insertion sequences using IScan. Nucleic Acids Res 35:5284–5293.

44. Touchon M, Rocha EPC. 2007. Causes of insertion sequences abundance in prokaryotic genomes. Mol Biol Evol 24:969–981.

45. Parkhill J, Wren BW, Thomson NR, Titball RW, Holden MTG, Prentice MB, Sebaihia M, James KD, Churcher C, Mungall KL, Baker S, Basham D, Bentley SD, Brooks K, Cerdeño-Tárraga AM, Chillingworth T, Cronin A, Davies RM, Davis P, Dougan G, Feltwell T, Hamlin N, Holroyd S, Jagels K, Karlyshev AV, Leather S, Moule S, Oyston PCF, Quail M, Rutherford K, Simmonds M, Skelton J, Stevens K, Whitehead S, Barrell BG. 2001. Genome sequence of Yersinia pestis, the causative agent of plague. Nature 413:523–527.

46. Zaghloul L, Tang C, Chin HY, Bek EJ, Lan R, Tanaka MM. 2007. The distribution of insertion sequences in the genome of Shigella flexneri strain 2457T. FEMS Microbiol Lett 277:197–204.

47. Bobay L-M, Ochman H. 2018. Factors driving effective population size and pan-genome evolution in bacteria. BMC Evol Biol 18:153.

