## Supplementary information - Figure S1 to S6 for "One Health genomics of *Acinetobacter baumannii* reveals sector-specific lineages and permeable ecological barriers" (from another version of this preprint: version 1, 2026-04-11)

**This document includes:**

- Figure S1. Geographic distribution of sampling sites and collected isolates of the Ab-One collection.

- Figure S2. Core-genome phylogeny and gene content similarity of *A. baumannii* clinical isolates of ST1639 and 2532 and belonging to the H clade.

- Figure S3. Core genome phylogeny of the 364 isolates of the Ab-One collection and of 471 representative genomes.

- Figure S4. Core-genome phylogeny and gene content similarity of *A. baumannii* cattle farm isolates belonging to the ST1027–1070 lineage.

- Figure S5. Core-genome phylogeny and gene content similarity of *A. baumannii* cattle farm isolates and selected isolates belonging to the ST927, 935 and 1013-2388 lineages.

- Figure S6. Number of prophage per genome in isolates of the three contexts.

**Separate files:**

table_S1.tsv : Ab-One isolates information - genome accession numbers - metadata

table_S2.tsv : Genome accession numbers and metadata of isolates from expanded phylogeny

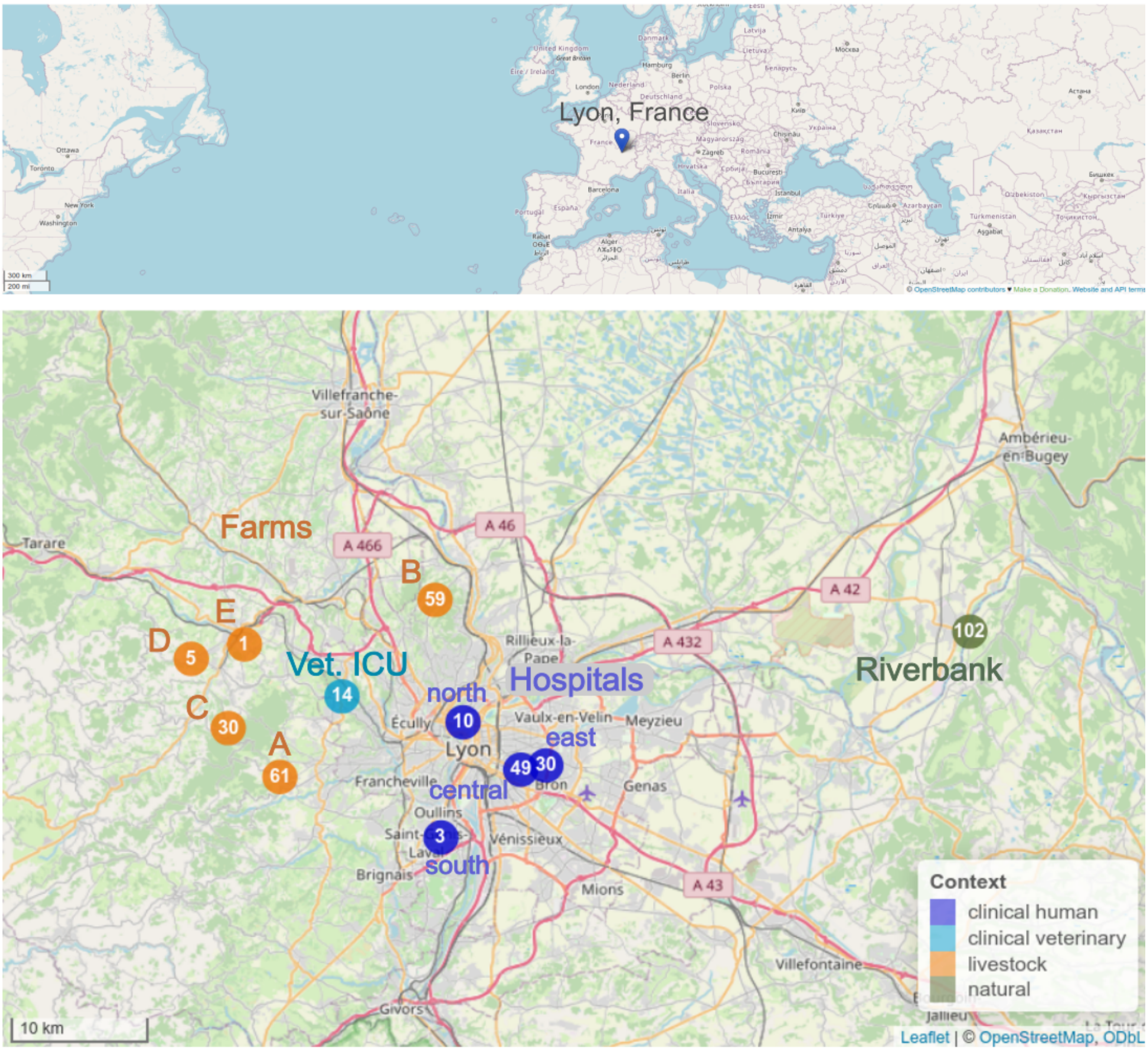


**Figure S1. Geographic distribution of sampling sites and collected isolates of the Ab-One collection.**

Top map shows the location of Lyon, France, within Europe. Bottom map provides a detailed map of the sampling area around the Lyon city, indicating the sites corresponding to each ecological context. Clinical human isolates (blue) were collected from the four civil hospitals sites (north, central, south, east); clinical veterinary isolates (turquoise) from the veterinary ICU (Vet. ICU); livestock isolates (orange) from cattle farms (A–E); and natural isolates (green) from the Rhône riverbank. Circles indicate individual sites, and numbers correspond to isolate counts per site. The base map was generated using OpenStreetMap data under the ODbL license.

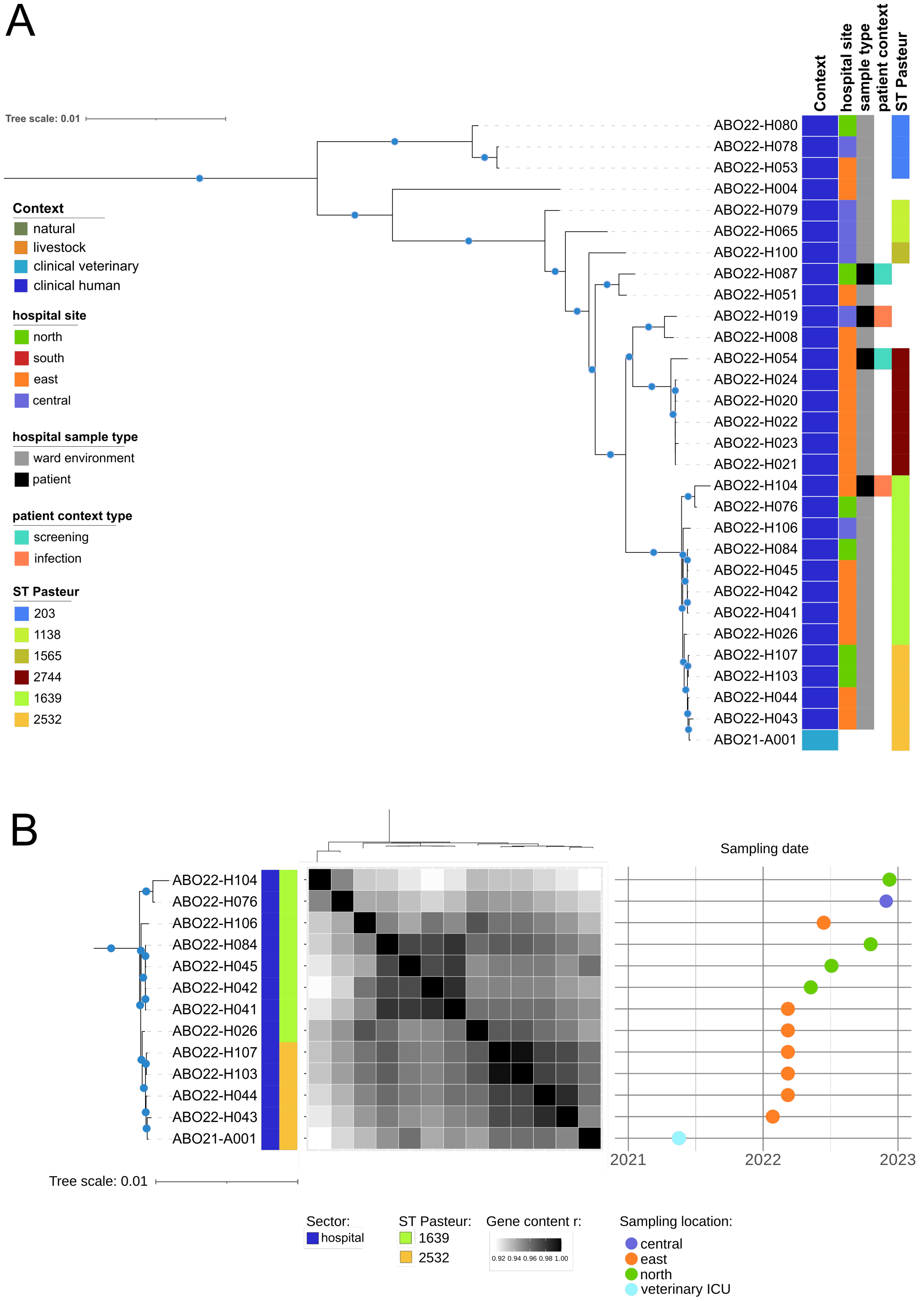


**Figure S2. Core-genome phylogeny and gene content similarity of *A. baumannii* clinical isolates of ST1639 and 2532 and belonging to the H clade.**

A. Phylogeny of the entire H clade. A and B, the phylogenetic tree (left; scale = 0.01 substitutions/site) shows clusters supported by bootstrap values ≥ 80 (blue dots). A and B, Adjacent color bars indicate sampling context. B. Focus on ST 1639 and 2532. The central heatmap displays pairwise gene-content similarity (r). To the right, the timeline indicates the sampling time of each isolate, with symbol colors denoting sampling location.

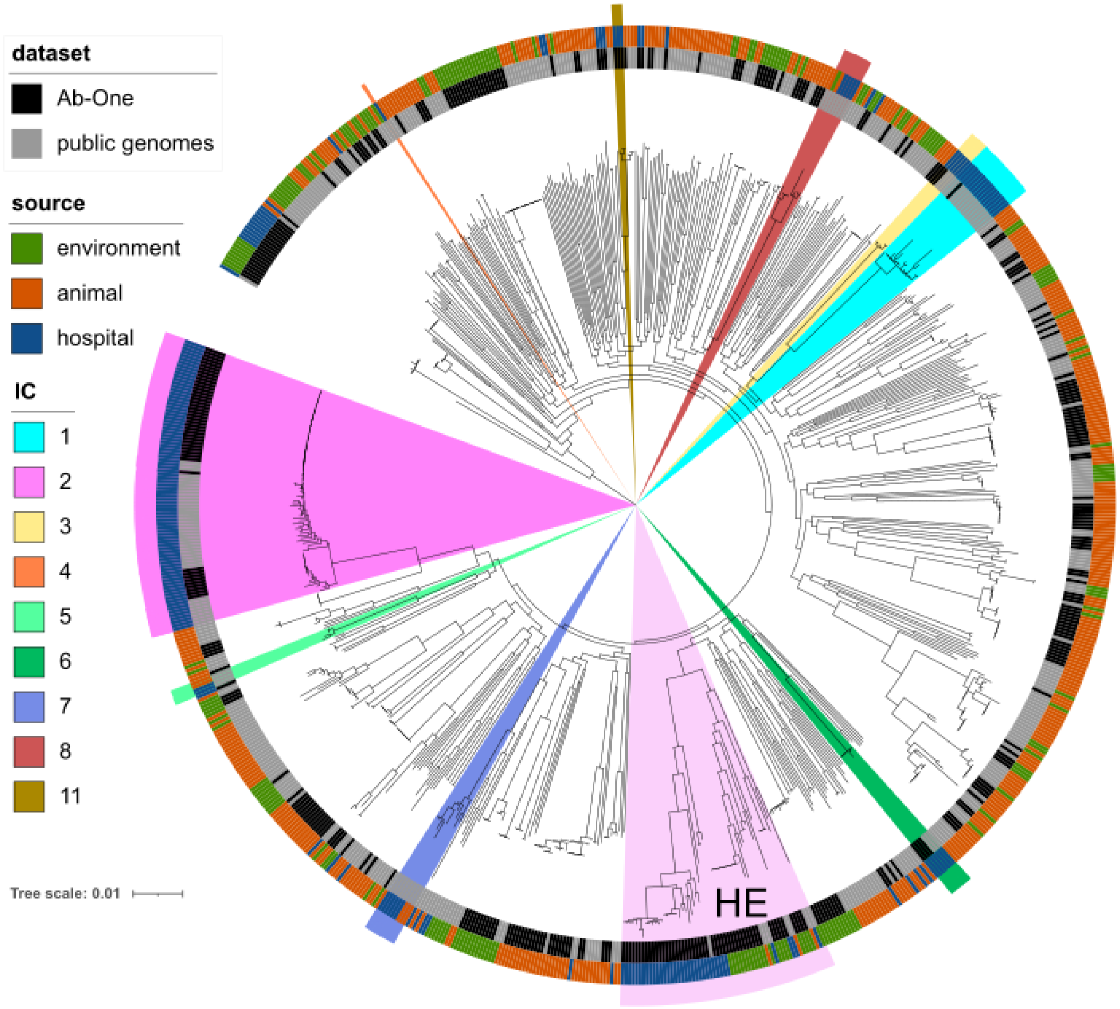


**Figure S3. Core genome phylogeny of the 364 isolates of the Ab-One collection and of 471 representative genomes.**

Maximum likelyhood phylogeny tree from the alignment of persistent genes (99% prevalence, 80% identity). The tree is rooted using five *A. nosocomialis* strains as an outgroup. Inner ring: source of the genomes. Middle ring: isolates are colored by the context of their isolation (hospital human and veterinary, dark blue ; livestock and wildlife, orange ; natural, green). Outer ring: international clone (IC). Tree scale is in substitution per site.

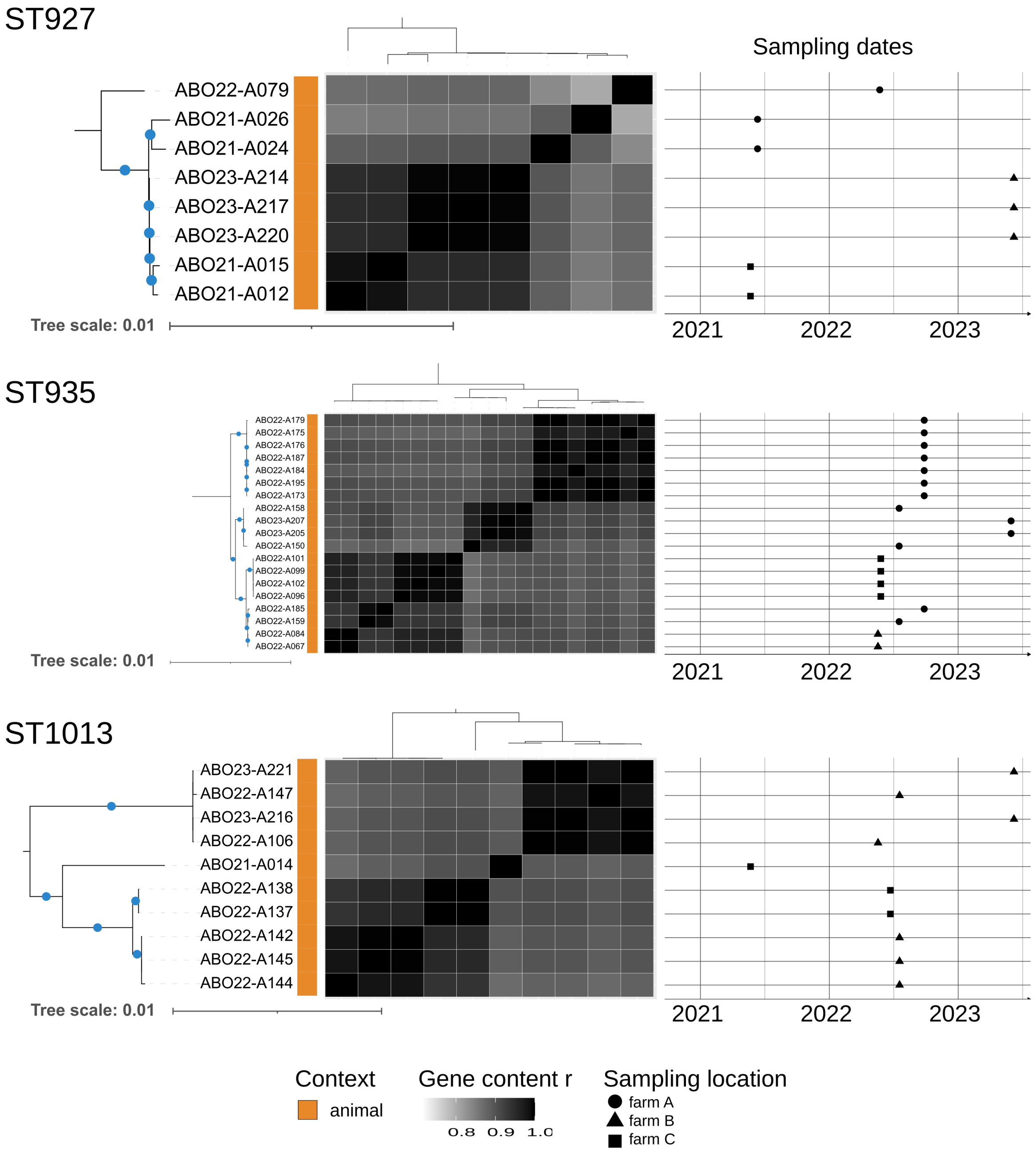


**Figure S4. Core-genome phylogeny and gene content similarity of *A. baumannii* cattle farm isolates belonging to the ST1027–1070 lineage.**
The phylogenetic tree (left; scale = 0.01 substitutions/site) shows clusters supported by bootstrap values ≥ 80 (blue dots). Adjacent color bars indicate sampling context. The central heatmap displays pairwise gene-content similarity (r). To the right, the timeline indicates the sampling year of each isolate, with symbols denoting sampling location.

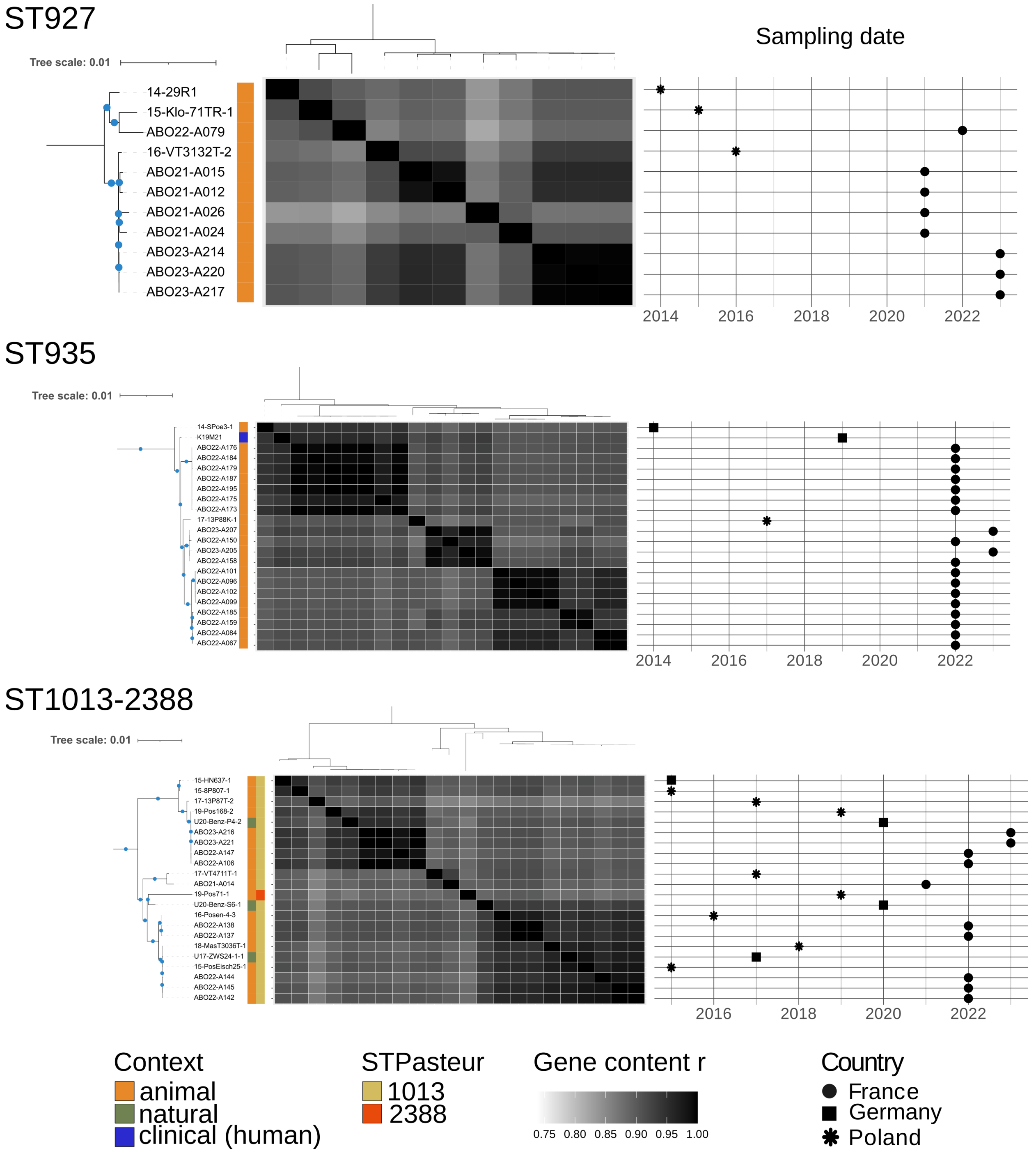


**Figure S5. Core-genome phylogeny and gene content similarity of *A. baumannii* cattle farm isolates and selected isolates belonging to the ST927, 935 and 1013-2388 lineages.**

The phylogenetic tree (left; scale = 0.01 substitutions/site) shows clusters supported by bootstrap values ≥ 80 (blue dots). Adjacent color bars indicate sampling context and ST type. The central heatmap displays pairwise gene-content similarity (r). To the right, the timeline indicates the sampling year of each isolate, with symbols denoting sampling country.

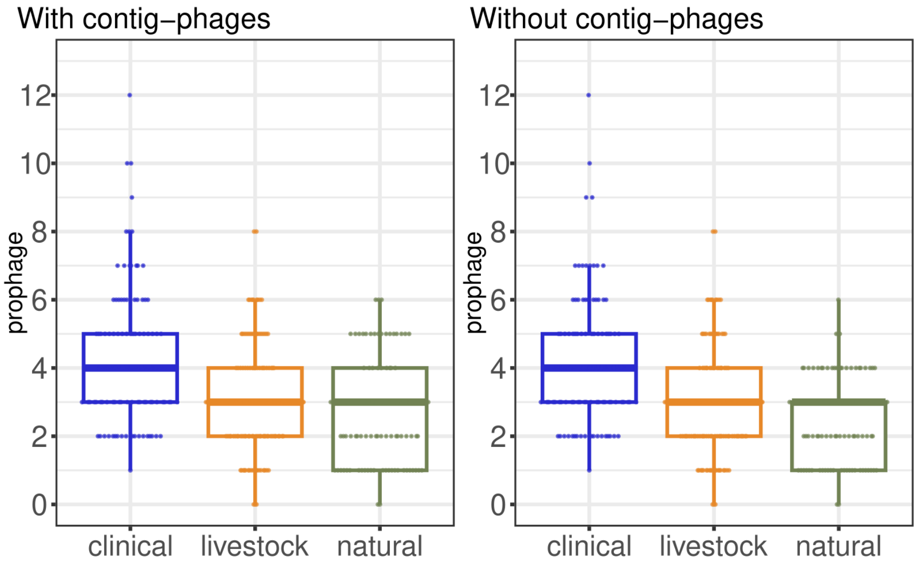


**Figure S6. Number of prophage per genome in isolates of the three contexts.**
Prophages were detected using geNomad in the complete genome (with contig-phage) or solely in the chromosome (without contig-phage).
